# Earlier Detection of Brain Injury Using Optical Brain Pulse Monitoring in Critically Ill Patients Following Cardiac Arrest

**DOI:** 10.1101/2025.02.18.25322327

**Authors:** Elliot J. Teo, Sigrid Petautschnig, Jack Hellerstedt, Sung W Chung, Andrew Udy, Paul Smith, Tim Haydon, Barry Dixon

## Abstract

**IMPORTANCE:** Point-of-care, non-invasive brain monitoring in critically ill patients following cardiac arrest could provide earlier detection of neurological injury and, when combined with earlier treatments, limit brain injury. Point-of-care monitoring could also enable better neuro-prognostication.

**OBJECTIVES:** The study assessed the time to detection of brain injury using optical brain pulse monitoring (OBPM) compared to routine brain monitoring. The association of OBPM signals with more severe forms of brain injury was also assessed.

**DESIGN:** Retrospective analysis of patients enrolled in an observational study.

**SETTING:** Critical care unit of a tertiary academic hospital.

**PARTICIPANTS:** Adult patients requiring mechanical ventilation in a critical care unit following a cardiac arrest.

**MAIN OUTCOMES AND MEASURES:** OBPM uses red and infrared light to capture brain pulse waveforms whose morphology reflects the relative arteriole and venous pressure levels driving microvascular blood flow in the brain. The OBPM sensors were placed bilaterally on the anterior temporal region of the scalp, over the middle cerebral artery territories. Time to brain injury detection was defined as the period from cardiac arrest to the first detection of brain injury by OBPM or routine monitoring.

**RESULTS:** Twelve patients were enrolled, three required veno-arterial extra-corporeal membrane oxygenator support. In-hospital mortality was 83% and eight patients developed global hypoxic-ischemic brain injury. The median time to detection of brain injury was 57 hours earlier using OBPM compared to routine monitoring (P < 0.01). In brain injured patients OBPM brain pulse morphologies changed over time and were often different between hemispheres, high amplitude respiratory waves were also present. Known poor prognostic brain pulse waveform morphologies were present in some patients with severe brain injury.

**CONCLUSIONS AND RELEVANCE:** OBPM detected brain injury earlier compared to routine brain monitoring. Earlier detection of neurological injury could improve patient outcomes through earlier treatment and better neuro-prognostication.

**KEY POINTS:** *Question:* Can point-of-care non-invasive optical brain pulse monitoring (OBPM) in critically ill patients following cardiac arrest provide earlier detection of brain injury compared to routine monitoring?

*Findings:* In this observational study of 12 patients the median time to detection of brain injury was 57 hours earlier using OBPM compared to routine monitoring.

*Meaning:* Earlier detection of brain injury could improve patient outcomes through earlier treatment and better neuro-prognostication.

## INTRODUCTION

Cardiac arrest requiring cardiopulmonary resuscitation affects around 26,000 people in Australia and 600,000 people in the United States annually. These patients are at high risk of neurological deterioration and require critical care unit management to optimise neurological outcomes [1–3]. Up to 10% of critical care admissions are cardiac arrest patients; however, survival of this patient cohort remains relatively low (<10%), and only 5% of patients have good neurological outcomes [1, 4, 5].

Common neurological complications following cardiac arrest, include cerebral oedema with raised intra-cranial pressure (ICP) and stroke [4, 6, 7]. Early detection of neurological deteriorations is an important goal to reduce death and disability, however this remains challenging [8–11]. Routine methods such as neurological examination or brain computerized tomography (CT) imaging often detects complications too late at an irreversible stage [12, 13]. Alternate approaches such as invasive brain monitoring, are complex and expensive and therefore infrequently used [14, 15].

Neuro-prognostication also remains challenging. Current assessments include clinical examination (including pupillary, corneal, and other reflexes), somatosensory evoked potentials, electroencephalogram (EEG), biomarkers, and brain imaging. However, each of these methods has limited accuracy [1, 16]. Better approaches to assess the severity of brain injury could enable earlier triage to assist families with decision-making in provision of complex, expensive and potentially futile therapies, such as extra-corporal membrane oxygenation (ECMO) or cardiac surgery [1, 17].

OBPM represents a novel, non-invasive approach to brain monitoring. OBPM detects microvascular cerebral blood flow responses following brain injury. OBPM uses red and infrared light to capture brain pulse waveform classes, whose morphology reflect the relative arteriole to venous pressure levels that drive microvascular blood flow [18]. Previous animal models and clinical work using OBPM has identified brain pulse classes including Arterial, Hybrid, Venous I, Venous II and Monotonous (Fig. 1) – that represent a continuum of blood flow states from normal (Arterial) to critically low blood flow (Monotonous). OBPM has been assessed in critically ill patients with acute brain injury [19–22]. A study in a human large vessel occlusion stroke and an ovine ischemic stroke model found the brain pulse classes were associated with the hypo-perfused stroke volume and also with long term outcomes including death [23, 24].

**Figure 1.**
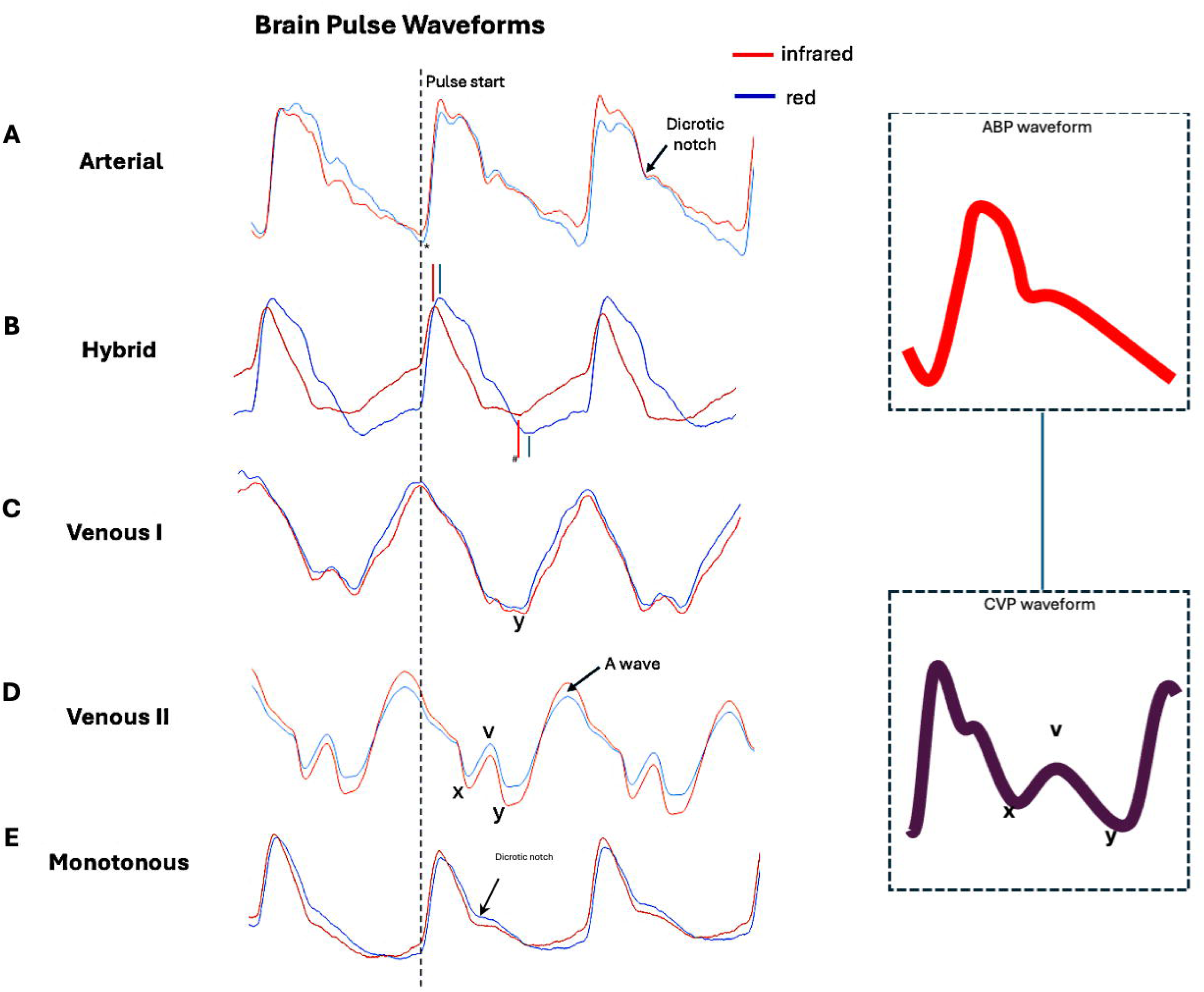
Brain pulse classes observed by OBPM. **A)** A normal **Arterial brain pulse** which has a shape similar shape to a normal arterial pressure pulse. B) The Hybrid brain pulse is characterized by distinct pulse shapes for the 940 nm and 660 nm wavelengths, due to their different responses to low blood oxygen levels. The pulse peaks * and troughs # of 660 nm are delayed relative to 940 nm. **C)** The **Venous I brain pulse** may have somewhat undifferentiated features. Consistent with similar cerebral arteriole and venous pressure levels throughout the cardiac cycle. The pulse may demonstrate more obvious central venous pressure features in the diastolic phase including V and Y waves. **D)** The **Venous II brain pulse** has clearer central venous pressure features such as the A, C, V, X and Y waves. The Venous II pulse is consistent with the venous pressure exceeding cerebral arteriole pressure throughout the cardiac cycle **E) Monotonous brain pulse** has absence of variation in shape between pulses and a low dicrotic notch. *Abbreviations: OBPM, Optical Brain Pulse Monitor;* Red brain pulse is 940 nm and blue brain pulse 660 nm.

This study assessed the time to brain injury detection using OBPM compared to routine monitoring and examined the possible association of OBPM signals with more severe forms of brain injury.

## METHODS

### Governance

St Vincent’s Hospital (Melbourne, Australia) Human Research Ethics Committee (HREC) granted ethics committee approval to the Transcutaneous Pulse Oximetry (T-POT) study [HREC 160/20], Project ID 63147, 14^th^ Dec 2021. Trial Registration: ACTRN12620000828921. https://www.anzctr.org.au/Trial/Registration/TrialReview.aspx?id=379639&isReview=true. Because participants lacked the capacity to consider participation at the time of eligibility, informed consent was obtained from their medical treatment decision maker. This study adhered to the Declaration of Helsinki and all methods were performed in accordance with the relevant guidelines and regulations of our hospital. The OBPM is a research device and is not yet cleared by the FDA for commercial use.

The inclusion criteria for the T-POT study were patients requiring admission to a critical care unit for mechanical ventilation following acute brain injury and or following a procedure with risk of brain injury and or cardiac or respiratory arrest, age > 18 years, and who would be considered for invasive brain monitoring. Patients were excluded if non-invasive brain monitoring was not possible due to a wound dressing, or other overlying skin or bone trauma. The current retrospective analysis represents the subgroup of cardiac arrest patients enrolled in the T-POT study. Patients with a history of previous brain injury were excluded from this analysis. This retrospective data analysis was approved through a protocol amendment.

### Optical brain pulse monitoring

The OBPM’s optical signal reflects pulsatile blood volume changes in the pial venules that lie on the cortical surface within the sub-arachnoid space. The pial venules relative blood volume is 4-fold higher than in the capillary beds of the cortex [25]. The pial venules act as a blood reservoir [26, 27]. Thus, a very large venous blood volume lies on the surface of the cortex, providing a strong optical signal. Functional MRI studies have demonstrated that the pial venous blood is dynamic in response to physiological changes in blood flow and dilate acutely following ischemic stroke [26, 28, 29]. The technological principles of OBPM and the methodology of clinical recordings have been previously reported [18, 24].

OBPM uses near infra-red (NIR) (660 nm) and red (940 nm) light sources to detect cardiac pulsations and respiratory waves arising from the brain. The features in these waveforms represent a combination of physiological responses including microvascular blood flow, blood oxygen and brain motion changes predominately associated with the cardiac and respiratory cycles [19, 20]. These features have been described previously and are summarised below [18, 24].

#### Brain cardiac pulse classes and relation to brain injury

We have identified distinct classes of OPBM brain pulses in normal and injured brains (Fig. 1). The Arterial brain pulse is seen in normal brains, while other brain pulses, including the Low compliance, Hybrid and Venous pulses are seen injured brains [18, 24]. The Low compliance brain pulse has similar features to invasive intracranial ICP monitoring waveforms with low brain compliance states or with raised ICP [20]. The Hybrid brain pulse is associated with low cerebral blood flow. The Venous I and Venous II brain pulses are both associated with very low or potentially retrograde cerebral blood flow [18, 24]. The Weak brain pulse also represents a very low cerebral blood flow state but does not have clear central venous features. The Monotonous pulse is likely associated with irreversible brain injury with infarction and absent cerebral arterial and venous blood flow [24].

#### Non-cardiac cycle related oscillations

Other classes of oscillations are present in injured brains that appear unrelated to the cardiac or respiratory cycles, such as Fast waves and Spindles which demonstrate high frequency oscillations (7-14 Hz) and also single Spikes [18]. We speculate that spasms (or fast oscillations) of cerebrovascular smooth muscle could be an underlying mechanism of Fast waves and Spikes (Fig. 1 Supplement) [18].

#### Respiratory waves

Intra-thoracic pressure oscillations associated with the respiratory cycle have marked influences on both brain venous flows and central spinal fluid (CSF) flows which in turn induce brain motion [30–34]. These responses give rise to respiratory waves detectable in the OPBM signal (Fig. 2 Supplement) [18]. In cases of brain injury, there may be notable differences in the amplitude of respiratory waves between hemispheres [18].

#### Optical intensity

The optical intensity assesses responses in 660 nm and 940 nm light absorption levels over longer time frames than the cardiac or respiratory cycles. The optical intensity is the raw unprocessed signal, reflecting light absorption by the skin and brain. Changes indicate slower variations in brain water or blood volume due to cerebral oedema, acute stroke or haemorrhage not related to the faster cardiac or respiratory frequencies. Slow oscillations (0.5–3 waves/minute) in cerebral blood flow or volume such as Lundberg B waves may be detected [22, 35, 36].

The OBPM is comprised of a roll-stand with an enclosure containing the Graphical User Interface (GUI) (a Tablet PC), a power supply for all components. The monitor’s light emitting diode (LED) and photodetector (PD) are controlled and processed by an Integrated Analog Front End circuit board, which digitizes the received signal from each sensor and sends the data stream to the Tablet PC. The PC receives the sensor data from the Processing Unit, and presents the data on a display, along with patient identifier data to the operator, via a custom software application. Data from the OBPM was captured at a rate of 500 Hz and stored for subsequent analysis. This analysis was conducted using Python v3.10, SciPy v1.10. The clean data were filtered to highlight specific frequency components relevant to physiological mechanisms for quantitative analyses.

### Data collection

Patient demographic data, physical examination findings, critical care and hospital length of stay, survival and other outcome data were collected from the hospital electronic patient record, as were the routine brain imaging and electroencephalograph (EEG) reports.

The OBPM sensors were placed bilaterally on the anterior temporal region of the scalp, over the middle cerebral artery (MCA) territories (Fig. 3 Supplement) [18]. Synchronous recordings were made with the routine intensive care monitoring outputs, including blood pressure, central venous pressure, ECG, end tidal CO_2_ and heart rate (Philips IntelliVue system using ICM+; Cambridge Enterprise, Cambridge, UK). Monitoring was undertaken Monday to Friday and was dependent on research staff availability. The OBPM was typically in place for approximately 1 – 12 hours.

### Data analysis

The brain pulse waveform class of the OBPM outputs were qualitatively assessed over both hemispheres. Brain Pulses were reviewed and classified based on their morphology into 6 distinct classes: 1) Arterial, 2) Low brain compliance, 3) Hybrid, 4) Venous I or II, 5) Weak and 6) Monotonous. The OBPM outputs were assessed by a single investigator (BD).

Increased respiratory wave amplitudes were present if the peak to trough amplitude of the respiratory wave in comparison to the peak to trough of the cardiac pulse was greater than 2:1. Optical intensity changes were defined as a > 2% change over 5 minutes in amplitude not clearly related with acute changes in blood pressure, body posture or movement artefact. Slow waves were defined as repeating oscillations in the optical intensity at a rate slower than the patient’s respiratory frequency [22].

The brain injury detection time was defined as the period since the cardiac arrest that brain injury was first detected by OBPM or routine monitoring. Brain injury was deemed present for OBPM by the presence of abnormal brain pulse classes, abnormal respiratory waveforms, fast waves, or acute changes in the optical intensity. Routine monitoring included CT or MRI to detect the presence of acute brain injury, EEG to show generalised epileptiform discharges or an isoelectric pattern, or clinical examination to demonstrate a major and consistent abnormality, such as myoclonus, hemiplegia, or fixed dilated pupils.

To assess if the OBPM was associated with the severity of brain injury, three categories of brain injury were defined. These from lower to more severe were; *1*) Regional injury, such as an ischemic stroke, documented on clinical examination or CT; or *2*) Global hypoxic-ischemic injury without brainstem coning, documented on CT; or *3*) Global hypoxic-ischemic injury documented on CT with coning (i.e., presence of fixed dilated pupils). Isoelectric EEG findings were used to determine the presence of global hypoxic-ischemic injury in one patient that did not have brain imaging. The prevalence of OBPM signals with these categories of brain injury was evaluated.

### Statistical Analysis

Graphs and statistical analyses were performed using GraphPad Prism version 9.4 (Graph Pad Inc, San Diego, USA). Aggregate patient demographic data is presented as median with interquartile range (IQR) or mean and standard deviation. Values of *P* < 0.05 were considered significant. Statistical analysis was undertaken by B.D a qualified biostatistician.

## Results

### Patient characteristics

Twelve subjects suffering cardiac arrest from the T-POT dataset were assessed (Table 1). The average age was 63 ± 10 years, three participants were female. The average time to return to spontaneous circulation (ROSC) after cardiac arrest was 30 ± 15 minutes. Ventricular tachycardia (VT) was the most common presenting rhythm, and cardiac aetiologies were the predominate underlying mechanism of cardiac arrest. Other aetiologies included sub-arachnoid haemorrhage (SAH), airway obstruction and pulmonary embolus. Three (23%) patients required support with veno-arterial (VA-ECMO). Ten (83%) of the 12 patients died during the hospital admission

**Table 1.** Patient characteristics.

|  |  |
| --- | --- |
|  | N = 12 |
| Age (years) | 63 ± 10 |
| Female | 3 (23%) |
| Time to return of spontaneous circulation | 30 ± 15 |
| Site of incident cardiac arrest |  |
| Out of hospital | 10 (83%) |
| In hospital | 2 (17%) |
| Cardiac rhythm at time of arrest |  |
| Ventricular tachycardia | 6 (50%) |
| Pulseless electrical activity | 5 (42%) |
| Asystole | 1 (8%) |
| Interventions |  |
| VA ECMO | 3 (25%) |
| CAGS or angiogram or PCI | 5 (42%) |
| Decompressive craniectomy | 1 (8%) |
| ICP monitoring | 2 (17%) |
| Active targeted temperature management | 12 (100%) |
| Presumed mechanism |  |
| Primary cardiac | 7 (58%) |
| Sub-arachnoid haemorrhage | 2 (17%) |
| Epiglottitis upper airway obstruction | 1 (8%) |
| Pulmonary embolus | 1 (8%) |
| Overdose | 1 (8%) |
| Brain injury documented on routine assessments | 12(100%) |
| Brain injury severity category |  |
| Regional brain injury | 4 (33%) |
| Diffuse hypoxic brain injury | 5 (42%) |
| Diffuse hypoxic brain injury with fixed dilated pupils | 3 (25%) |
| Outcome |  |
| Hospital length of stay (days) | 10 ± 7 |
| CCU length of stay (days) | 6 ± 5 |
| Died | 10 (83%) |
| Transfer to another hospital | 2 (17%) |
| WLST/Palliation | 10 (83%) |

**Legend:** WLST = withdrawal of life supporting treatments, VA-ECMO = venous arterial extra-corporeal membrane oxygenation, CAGS; coronary artery bypass grafting, ICP: intra-cranial pressure, CCU, critical care unit

### Severity of brain injury

All 12 patients had evidence of brain injury on routine monitoring at some point during their critical care admission. The severity of brain injury varied. Three patients (33%) had a regional brain injury with a focal stroke on CT or clinical examination, five patients (42%) developed global hypoxic-ischemic brain injury and three (23%) developed global hypoxic brain injury with coning. (Table 1 supplement).

### Detection of brain injury

The median time from cardiac arrest to commencement of OBPM was 20 (IQR 9 – 27) hours. The duration of monitoring increased over the course of the study, as hospital staff gained experienced with the monitor. The mean duration of OBPM monitoring per patient was 13 ± 16 hours. The days of monitoring ranged from the day of injury (day 0) to day 7.

The time to detection of brain injury was assessable in eight patients. Four patients were not assessable as brain injury was documented by routine monitoring before OBPM was commenced; two patients presented with SAH and two with evidence of global hypoxic-ischemic on early CT. In the eight assessable patients, the median time to detection of brain injury was 77 (IQR 37 - 122) hours with routine monitoring, whilst the time was 20 (IQR 9 – 27) hours for OBPM (P < 0.01; Mann Whitney test, Fig. 2).

**Figure 2.**
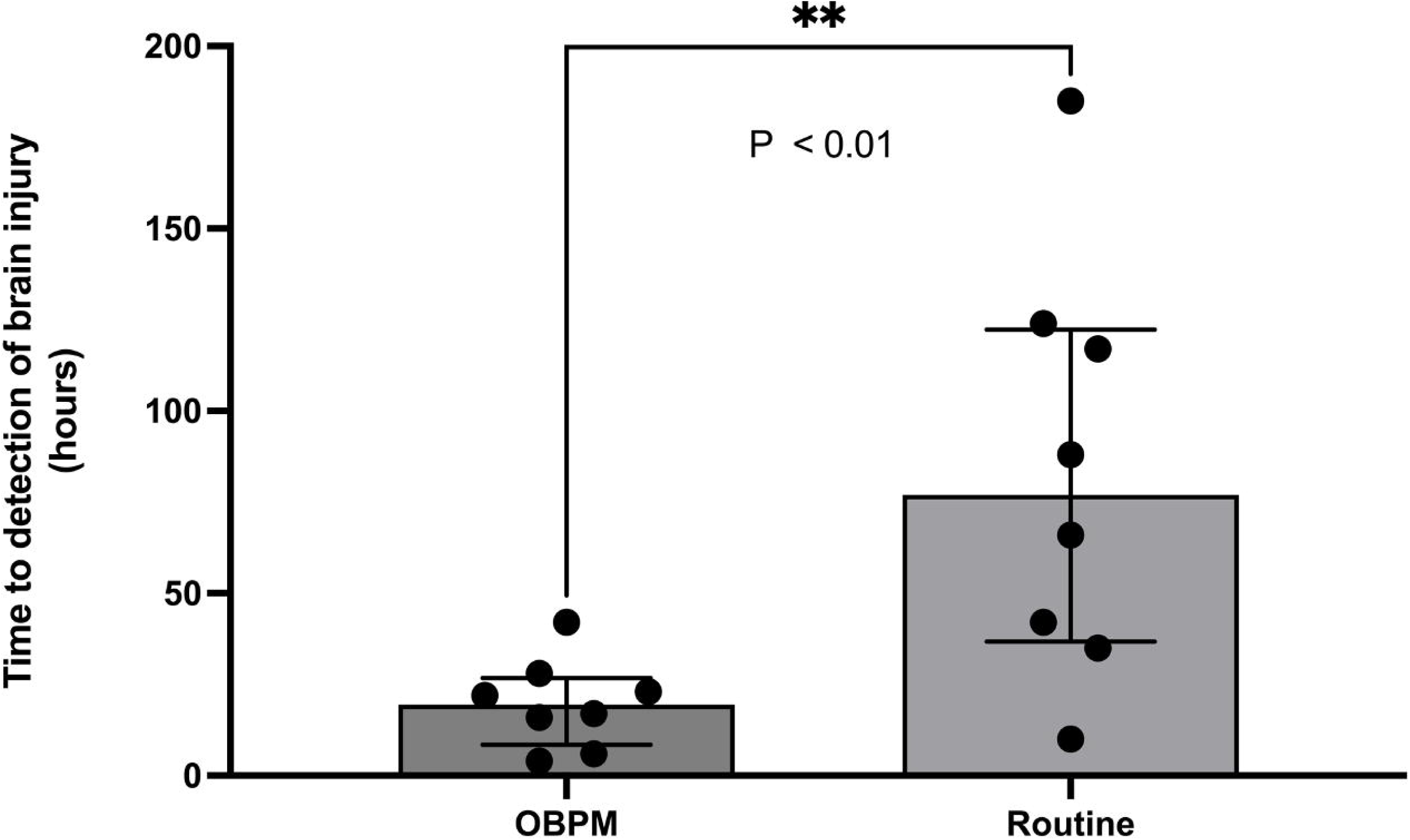
The optical brain pulse monitor (OBPM) detected brain injury 57 hours earlier compared to routine monitoring; 20 (IQR 9 – 27) as compared to 77 (IQR 37 - 122) hours (P < 0.01). The time to detection represents the time since the onset of the cardiac arrest. *Abbreviations; IQR, Inter-quartile range*

Routine monitoring documented brain injury the following ways, CT reporting (n = 4), Isoelectric EEG (n=1), clinical detection of fixed dilated pupils (n=2) and clinical examination finding of hemiplegia (n=1). Of interest, early CT brain imaging was normal in four patients who subsequently developed routine monitoring evidence of brain injury on later imaging.

### Pattern of abnormal OBPM signals in relation to brain injury severity

The pattern of abnormal OBPM signals, including the brain pulses classes, high respiratory amplitude, fast waves and optical intensity changes were assessed for each of the categories of brain injury. Abnormal OBPM signals were present in all 12 patients. There was variability in the prevalence of abnormal OBPM signals for each of the severity categories of brain injury (Fig. 4 Supplement). High amplitude respiratory waves were present in all patients. The average number of distinct types of abnormal OBPM signals was 3.0 for patients with regional brain injury, 3.8 for global hypoxic-ischemic injury and 5.3 for global hypoxic-ischemic injury with brainstem coning (P = 0.3, ANOVA). Known poor prognostic brain pulse classes such as Venous II and Monotonous were present in two patients, both subsequently developed global hypoxic-ischemic injury on CT with brainstem coning (Fig. 3)

**Figure 3 A.**
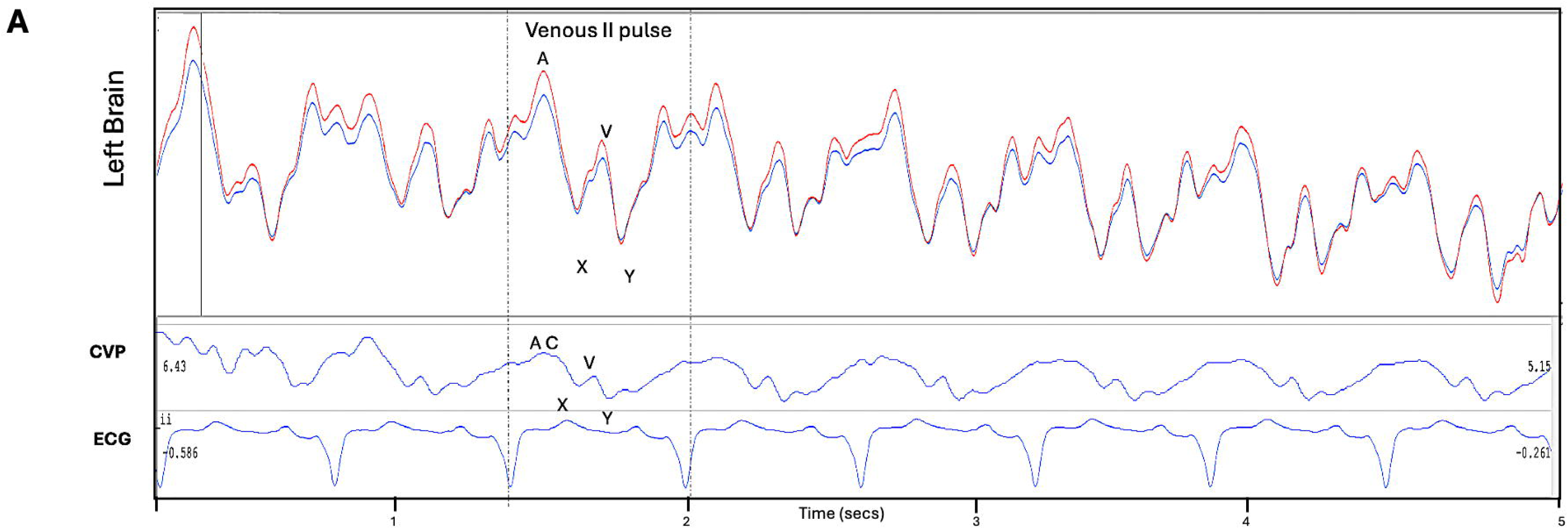
**Venous II brain pulse** present over left brain in a patient on extra corporeal veno-arterial membrane oxygenator following out of hospital cardiac arrest (the right brain sensor was disconnected). The concurrent central venous pressure (CVP) trace is demonstrated. The venous circulation A, C, X, V, Y waves are labelled. A **Venous II brain pulse** suggests very low or retrograde cerebral flow and low brain oxygen levels in the monitored region of the brain and suggests a poor prognosis. The patient developed global hypoxic injury and died. Red brain pulse is 940 nm and blue brain pulse 660 nm.

**Figure 3 B.**
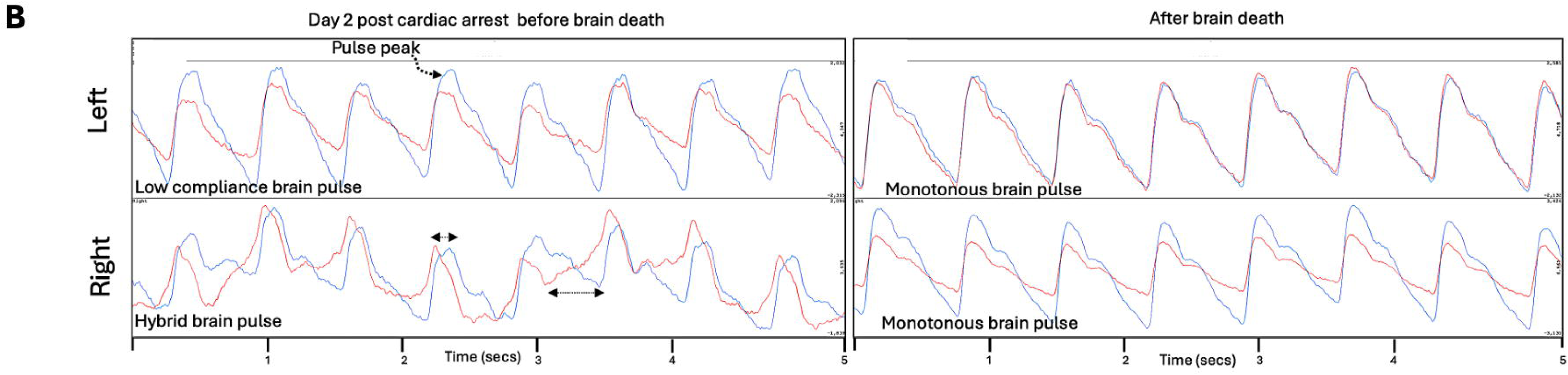
Another patient on day 2 following out of hospital cardiac arrest (left panel) and following clinical evidence of brain death **Monotonous brain pulses** developed (right panel). The patient developed cerebral oedema with fixed dilated pupils on day 3 and was confirmed on examination to be brain dead. On day 2 the left brain demonstrated **Low brain compliance pulse** features including a late pulse peak and high dicrotic notch. The right brain demonstrates a **Hybrid brain pulse** with marked asymmetry of 660 and 940 nm pulse shapes, the pulse peaks and troughs of 660 nm are delayed relative to 940 nm (arrows). These features suggest swelling and cerebral oedema of the left brain and reduced cerebral blood flow of the right brain. On day 6 following brain death the left and right brains both demonstrate **Monotonous brain pulses** with absence of variability. In addition, there is strong symmetry of the 660 and 940 nm pulse shapes. The **Monotonous brain pulses** and lack of variation suggest absent cerebral blood flow. We speculate this pattern may be due to CSF induced flow or pressure waves transmitted from the spinal fluid induced by spinal arterial pressure pulses. *Abbreviations* CVP: central venous pressure, ECG: electrocardiogram. A figure similar to 3A has previously been published in Med Devices, 2024 Dec 11:17:491-511.

### OBPM pulse classes and other signals changed over time with brain injury

In all 12 patients the brain pulse OBPM signals changed frequently over the hours and days of the study. In one patient a change occurred immediately following cardioversion to sinus rhythm and was associated with an acute neurological deterioration. The optical intensity fell immediately in the left hemisphere following cardioversion from prolonged ventricular tachycardia (VT) to sinus rhythm in a patient on VA ECMO (Fig. 4A). An hour later, the patient then developed high amplitude slow waves (possibly Lundberg C waves) in the right hemisphere (Fig. 4B, right panel) and 7 hours later the patient developed fixed dilated pupils with coning. A subsequent CT demonstrated global hypoxic-ischemic injury and the patient died.

**Figure 4A.**
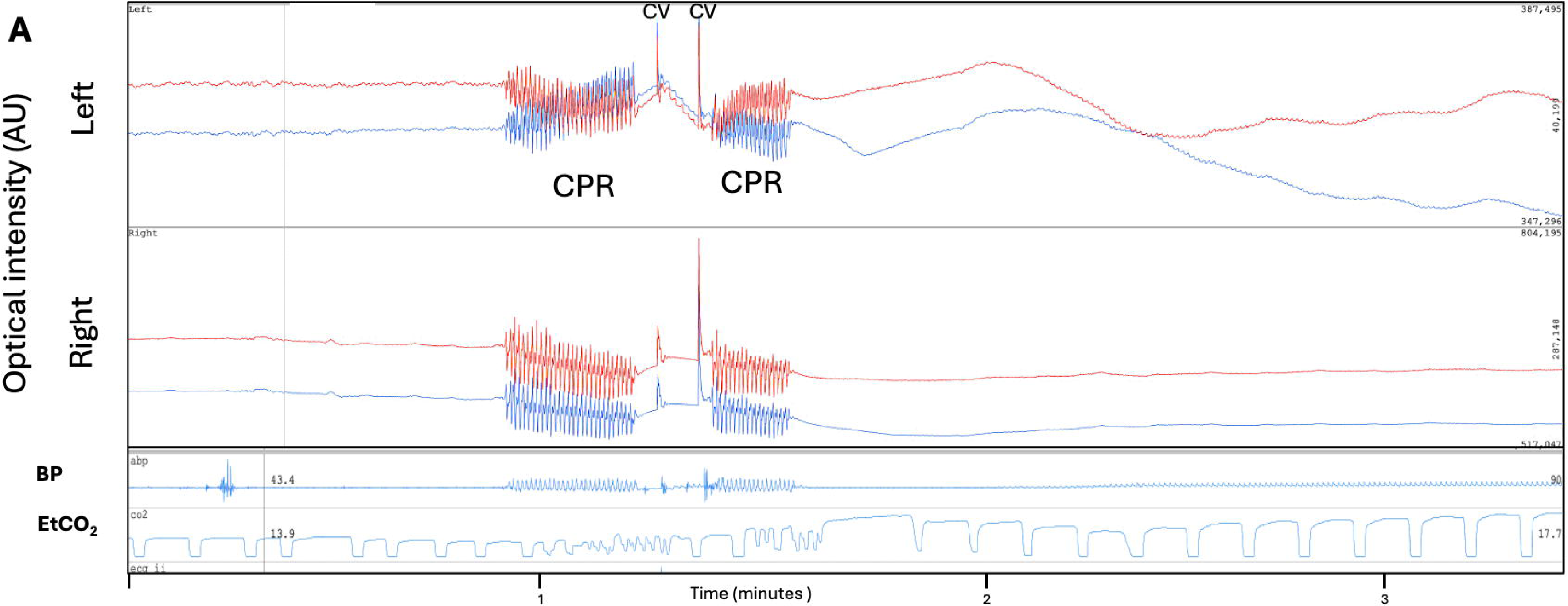
**Brain optical intensity (OI)** levels for 940 nm (red) and 660 nm (blue) before and following cardioversion (CV) with cardiopulmonary resuscitation (CPR) to revert prolonged ventricular tachycardia to sinus rhythm in a patient on veno-arterial a extra corporeal membrane oxygenator (ECMO). Acute changes in the OI developed over minutes in the left-brain following cardioversion. The patient developed fixed dilated pupils 11 hours after sinus rhythm was restored. We speculate that a left ventricular clot may have embolised to the brain following cardioversion to sinus rhythm. A subsequent CT demonstrated diffuse cerebral oedema; whether a stroke occurred earlier as a precipitant or not could not therefore be confirmed.

**Figure 4B.**
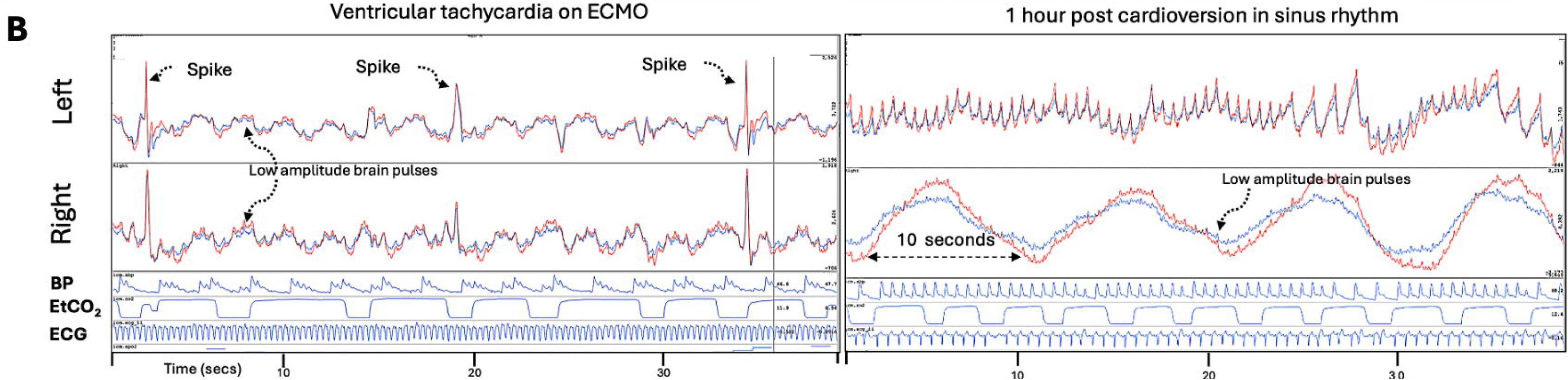
Same patient demonstrating the brain pulse signals during ventricular tachycardia (left panel) and following cardioversion (CV) while in sinus rhythm (right panel). **Left panel** demonstrates very low amplitude brain pulses with frequent Spikes. **Right panel.** An hour following cardioversion the right hemisphere developed slow oscillations, potentially representing Lundberg C waves and consistent with developing brain injury. *Abbreviations* BP: blood pressure, EtCO_2_ : End-tidal CO_2_, ECG: electrocardiogram, AU: arbitrary units. A figure similar to 4A has previously been published in Med Devices, 2024 Dec 11:17:491-511.

### OBPM brain pulse classes were predominantly different between hemispheres in brain injury

Over the monitoring periods the brain pulse classes were different between hemispheres 70% of the time. Fig. 5 provides one example with a Hybrid brain pulse on the left brain, suggestive of low cerebral blood flow and a Low compliance brain pulse on the right brain, suggestive of brain swelling. Other examples are included in the supplement see Figs. 5 and 6.

**Figure 5.**
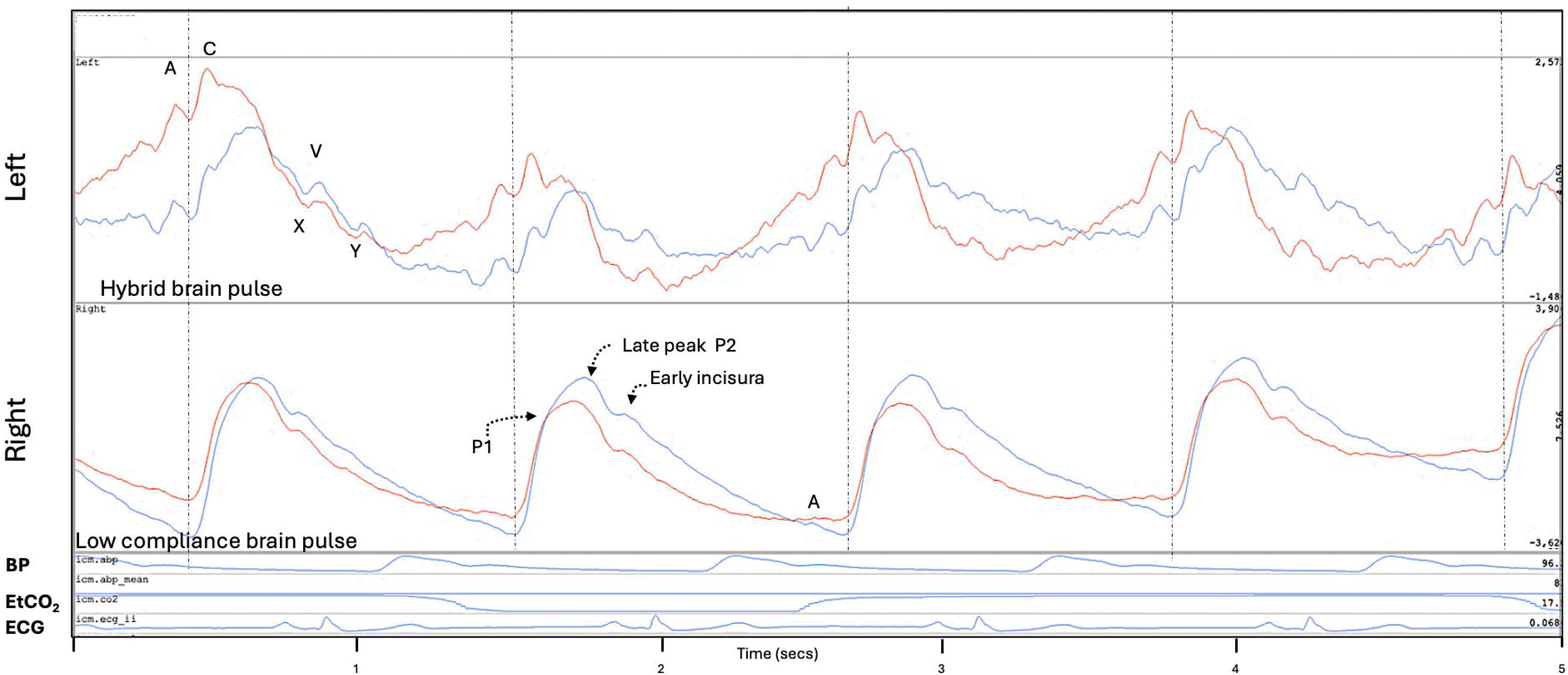
Left brain **Hybrid brain pulse** is characterized by distinct shapes for the 940 nm and 660 nm wavelengths, due to their different responses to low blood oxygen levels. The pulse peaks and troughs of 660 nm are delayed relative to 940 nm and subtle venous circulation features A, C, X, V and Y waves may be present. These features suggest low cerebral blood flow blood oxygen levels. The right brain has a **Low compliance brain pulse** has features like an invasively measured intra-cranial pressure pulse with low brain compliance or high ICP including a delayed time to P2 and high P2/P1 ratio, subtle A waves may be present. These findings are consistent with right sided cerebral oedema or brain swelling with low cerebral blood flow oh the left. Red brain pulse is 940 nm and blue brain pulse 660 nm. *Abbreviations* BP: blood pressure, EtCO_2_: End-tidal CO_2_, ECG: electrocardiogram.

### High amplitude respiratory waves

High amplitude respiratory waves were present in all patients. In three patients, a distinctive asymmetrical hemispheric pattern of respiratory and brain pulse changes was noted. The respiratory wave amplitude was very high on the left hemisphere, whereas the right hemisphere the respiratory amplitude was normal with a Low compliance brain pulse (Supplement Fig. 7).

## Discussion

In this exploratory study, we found of a novel, point-of-care brain monitor (OBPM) identified the presence of brain injury 57 hours earlier than routine monitoring in patients following cardiac arrest.

### Early detection of brain injury

Early detection of neurological deterioration is seen as an important goal to reduce death and disability [8–11] and is recommended by expert panels, including the Australian Trauma Guidelines and The International Consensus Conference on Monitoring in Neurocritical Care [37–39]. Earlier detection of neurological deterioration allows earlier treatments to prevent irreversible brain injury. Treatments include altered blood pressure targets, blood transfusion, increased sedation, modified ventilation modes or earlier brain imaging to identify complications that could benefit from urgent invasive treatments, including decompressive craniectomy or mechanical thrombectomy [1, 40].

The large time difference of 57 hours between OBPM and routine monitoring may appear surprising, however patients managed in critical care units following cardiac arrest are commonly actively cooled to protect their brain for 72 hours [41]. While cooled, patients are heavily sedated and maybe paralysed and, consequently, adequate neurological examination is not possible. Furthermore, CT in the first 24 hours is poor at detecting brain injury and is therefore generally not performed before 72 hours [13].The median time from cardiac arrest to OBPM placement was 20 hours which suggests our results could be improved by commencing monitoring earlier.

Our findings suggest OBPM captures cerebrovascular responses not detected by CT imaging. Early CT brain imaging was normal in four patients who subsequently developed evidence of brain injury. In all these cases OBPM demonstrated abnormal findings before routine monitoring. This highlights the distinct physiological information captured by OBPM including pial venous blood oxygen levels, cerebral blood flow and brain motion changes associated with the cardiac and respiratory cycles [19–21].

### Relationship of OBPM signals with brain injury severity

While there was variation in the prevalence of distinct brain pulse classes and other signals amongst the categories of severity of brain injury, there was no statistical difference found. The small sample size of our study likely meant that it was underpowered to assess for an association between OBPM abnormal signal classes and worse brain injury with poor prognosis.

Brain pulse classes such as Venous II and Monotonous were however present. These have been demonstrated in an earlier study of patients presenting with large strokes to be associated with worse long term outcomes including death [24]. One patient with a persistent Venous II pulse class subsequently developed global hypoxic ischaemic injury with fixed dilated pupils and died. In this patient the central venous pressure trace was simultaneously recorded. The pulse shapes were remarkably similar for the brain pulse and the central venous pressure wave forms, suggesting a venous circulation origin (Fig. 3A). The presence of the Venous II pulse is consistent with cerebral venous pressure exceeding cerebral arteriole pressure throughout the cardiac cycle, which would likely result in retrograde or reversed cerebral blood flow. Reversal of blood flow in cerebral veins has previously been demonstrated in critically ill patients with severe brain injury using transcranial doppler [42, 43]. Reversal of blood flow has also been demonstrated in cortical arterioles, capillary beds and venules in animal models of stroke [44–47].The prolonged presence of the Venous II pulse is therefore likely to be associated with a poor prognosis.

In another patient in which brain death had occurred the brain pulse developed Monotonous features over both hemispheres with a remarkable absence of pulse variability over time and high symmetry of the 660 and 940 nm pulse shapes (Fig. 3B). The Monotonous brain pulse has an arterial pressure waveform shape but has some atypical features including a low dicrotic notch and an absence of waveform shape variation over time. Both the low dicrotic notch and the absence of physiological variation over time are consistent with absent cerebral blood flow. Similar characteristics have been demonstrated in intra-cranial pressure waveforms in patients with established brain death [48, 49]. The low dicrotic notch is consistent with absence of a reflected pressure wave back from the brain arterioles [50]. The mechanism of this pulse may represent a transmitted arterial pressure wave through the brain tissues rather than through the brain’s circulation. The Monotonous brain pulse suggests irreversible brain injury with infarction with absent arterial and venous blood flow [24].

### Respiratory waves and brain injury

We found high respiratory wave amplitudes were common in brain injury. An interesting asymmetrical pattern of change was observed in 3 patients where one hemisphere had a high respiratory amplitude while the contralateral hemisphere it was normal and associated with a Low compliance brain pulse (Fig. 7 Supplement). These features are consistent swelling or cerebral oedema of the Low compliance brain pulse side and may be associated with mid-line shift [18]. The swelling or increased brain volume may divert the cranial respiratory flow of CSF to the more compliant contralateral hemisphere resulting in an exaggerated respiratory motion [51–54]. Other studies using MRI following brain injury also found large unilateral hemispheric respiratory cycle brain motion abnormalities associated with disturbed CSF flows in the lateral ventricles [51, 52]. Similar responses were found with decompressive craniotomy [55]. Lundberg also found marked increases in ICP respiratory amplitudes following traumatic brain injury (TBI) [56].

### Hemispheric differences and temporal changes in optical signals with brain injury

The OBPM brain pulses and other optical signals were usually different between hemispheres in brain injured patients. The signal pattern differed between hemispheres 70% of the time. EEG studies in brain injured patients found that the Bi-spectral index, which assesses the degree of asymmetry of the EEG signal was the strongest predictor of poor neurological outcomes in TBI patients [57]. Our signal outputs were also dynamic changing over hours and days of the study. This is consistent with the evolving brain injury and inflammatory responses seen on brain CT and MRI scans, EEG and pupillometry following cardiac arrest [16, 58, 59].

### ECMO and potential detection of an acute neurological complication

Three patients required VA ECMO for haemodynamic support following cardiac arrest. Despite the reduced cardiac stroke volume brain cardiac pulses and respiratory waves remained present in the OBPM signals. In one case we documented acute changes in the left hemisphere optical intensity developed immediately following cardioversion from VT to sinus rhythm (Fig 4A). Over the next hour the patient developed slow (possibly Lundberg C) waves with a frequency of 6 per minute over the contralateral right hemisphere (Fig. 4B, right panel). Seven hours later the patient had fixed dilated pupils. We speculate that a cardiac clot may have embolised to the brain following cardioversion to sinus rhythm. A subsequent CT demonstrated global hypoxic-ischemic injury, whether a stroke occurred earlier as a precipitant or not could not be confirmed. Acute brain injury occurs in 8% of adults on VA-ECMO, due to intracranial haemorrhage, stroke, and hypoxic-ischemic brain injury [60, 61]. A method to detect this earlier and provide treatments before irreversible injury could improve patient outcomes and provide earlier neuro-prognostication.

Other NIR devices have been used to assess evidence of brain injury and to influence treatments without strong evidence of efficacy to date. A recent large, randomised study using the INVOS cerebral oximeter in neonates found no benefit [62]. A study of the INVOS cerebral oximeter in patients with TBI demonstrated a low accuracy at detecting deteriorations associated with cerebral deoxygenation [63]. Two subsequent larger studies of out-of-hospital cardiac arrest (OHCA) patients also did not find a clinically significant association between post-cardiac arrest rSO_2_ with neurological outcomes [64, 65].

### Limitations

A limitation was the small size of this study; therefore, the analyses were exploratory in nature and a larger study is required to confirm the current findings.

The duration and timing of OBPM monitoring varied amongst the patients. Monitoring the brain responses at consistent time points and for longer periods would have provided better understanding of the pathophysiological responses following cardiac arrest.

The skull has a range of muscles attached, such as the temporalis muscle (which the sensor sits over), chewing, swallowing can cause movement of the sensor and therefore motion artefact. Other causes of sensor movement, such as change in body position, shivering, coughing, head movement or sensor contact with the patients’ bedding can also cause artefacts, as tension or movement on the cables attached to the sensors. These artefacts could be mistaken for a OBPM signal.

The waveform patterns were assessed by a single author (BD), future studies would benefit from blinded assessments by multiple assessors. The study was undertaken in a single critical care unit in Australia whose practices may not be different to other international centres [66]. The categories of severity of brain injury were chosen based on the narrow range available in a retrospective cohort of patients with a very low survival rate. A future larger study assessing longer term functional outcomes would be of value.

## Conclusions

OBPM detected brain injury earlier compared to routine brain monitoring. Earlier detection of neurological deteriorations could improve patient outcomes through earlier treatment. Improved neuro-prognostication could also assist in providing better patient centred care.

## Data Availability

All data produced in the present work are contained in the manuscript

## Acknowledgments

We would like to acknowledge Murray Worner and Leigh Baker of the Department of Medical Imaging and Physics for assistance in the study.

## Author contribution

All authors made a significant contribution to the work reported, whether that is in the conception, study design, execution, acquisition of data, analysis and interpretation, or in all these areas; took part in drafting, revising or critically reviewing the article; gave final approval of the version to be published; have agreed on the journal to which the article has been submitted; and agree to be accountable for all aspects of the work. Specific contributions included but were not limited to the following: BD conceived the study. BD designed the human study protocol. SP, BD, and EJT collected data. BD, SP, SWC, JH and EJT analysed data and interpreted the results. BD wrote the first manuscript draft.

