## Supplement for "Earlier Detection of Brain Injury Using Optical Brain Pulse Monitoring in Critically Ill Patients Following Cardiac Arrest"

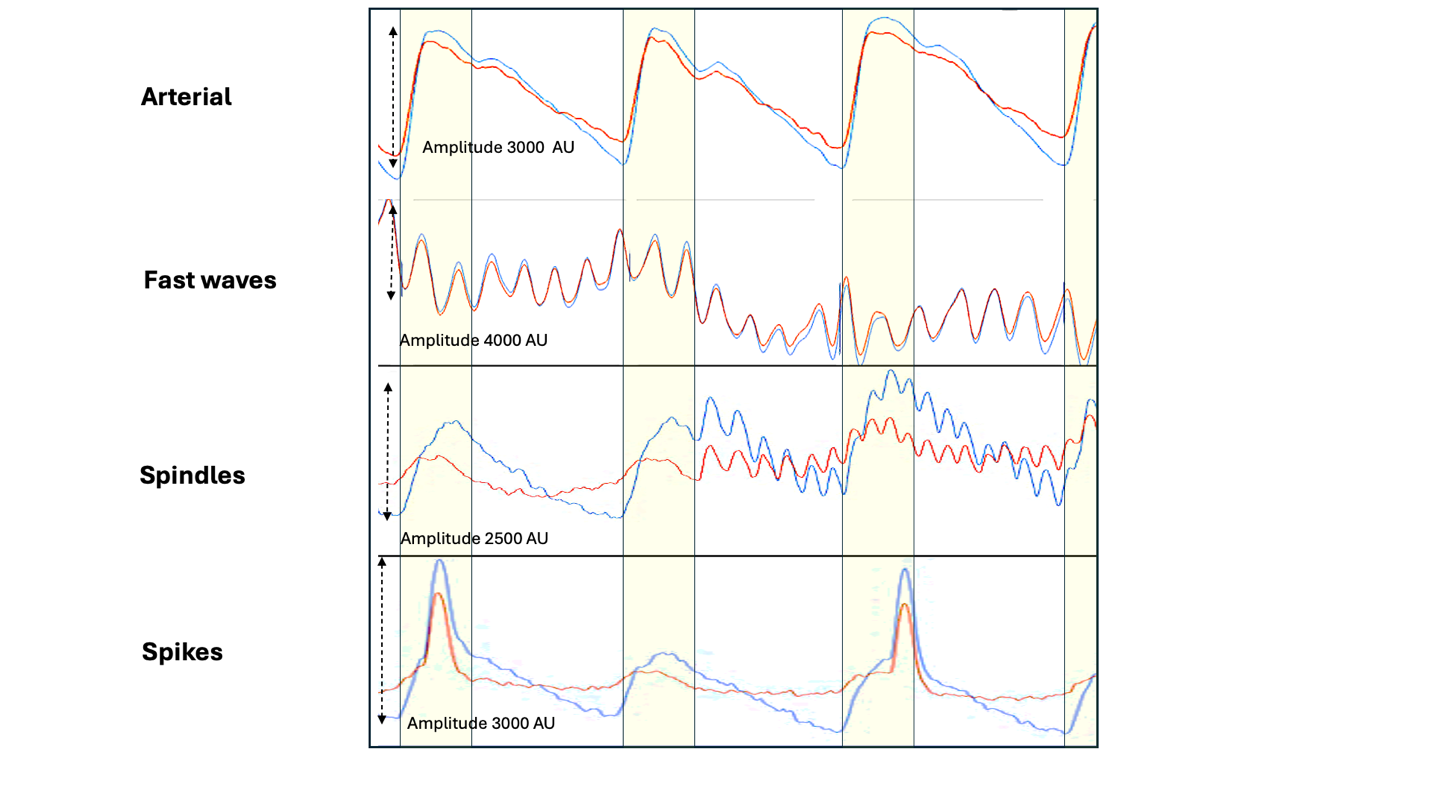

**Figure 1.** Demonstrates a normal **Arterial** brain pulse and brain injured waves**; Fast waves** oscillate at high frequencies (8-14 Hz). The amplitude is typically high > 2000 AU with symmetry of 940 and 660 nm oscillations throughout the cardiac cycle. **Spindles** also oscillate at high frequencies (10 -14 Hz), but are typically brief, just a few heart beats in duration and the amplitude is lower than Fast waves. **Spikes** are typically single brief waves. They are usually independent of the cardiac cycle. Red brain pulse is 940 nm and blue brain pulse 660 nm. The yellow area represents the systolic phase of the cardiac cycle and the white the diastolic phase. *Abbreviations:* AU: arbitrary units. This Fig has previously been published in Med Devices, 2024 Dec 11:17:491-511.

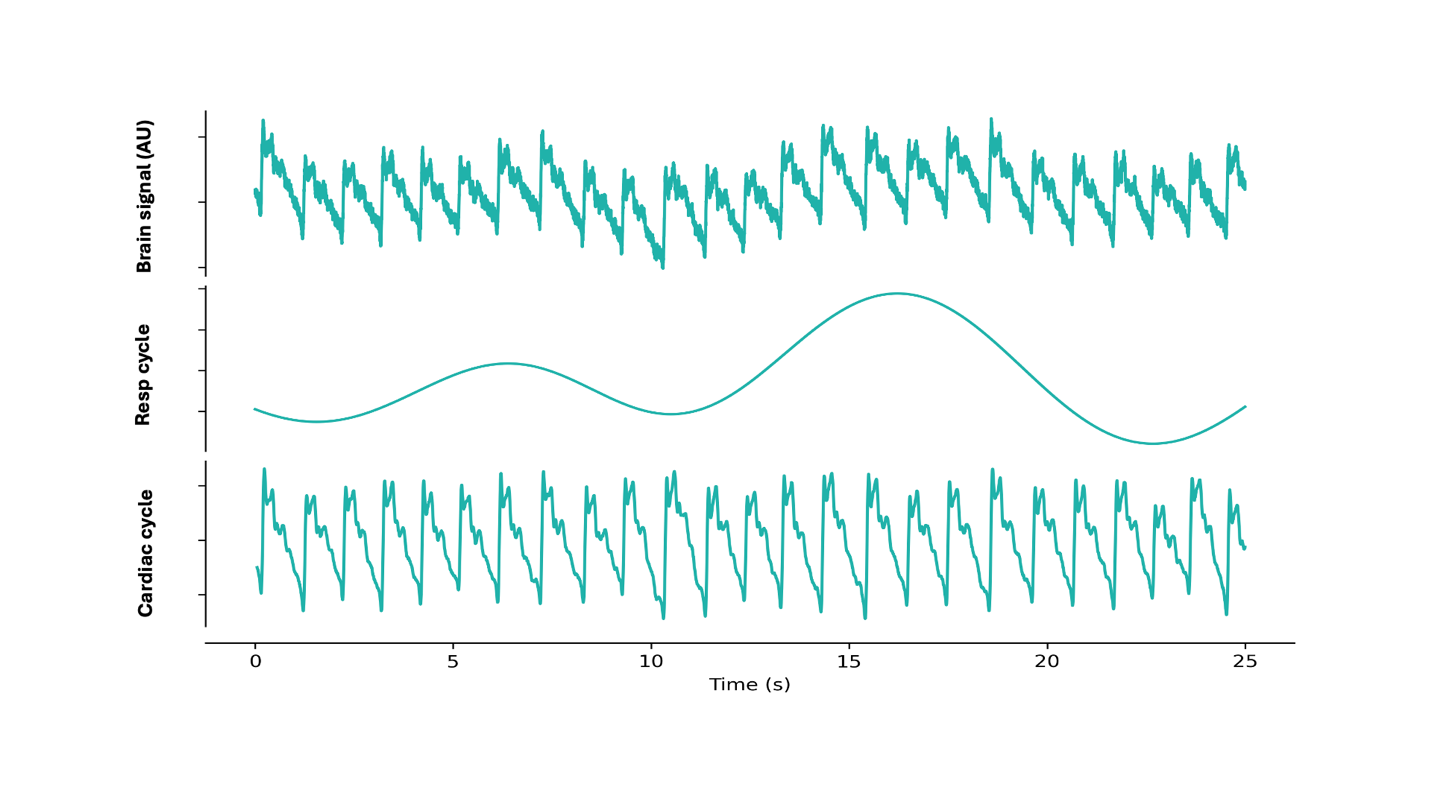

**Figure 2.** Separation of the raw optical brain pulse signal (upper panel) with signal processing into its components, the respiratory cycle wave (middle panel) and cardiac cycle pulse (lower panel). Resp: Respiratory, AU: arbitrary units. This Fig has previously been published in Med Devices, 2024 Dec 11:17:491-511.

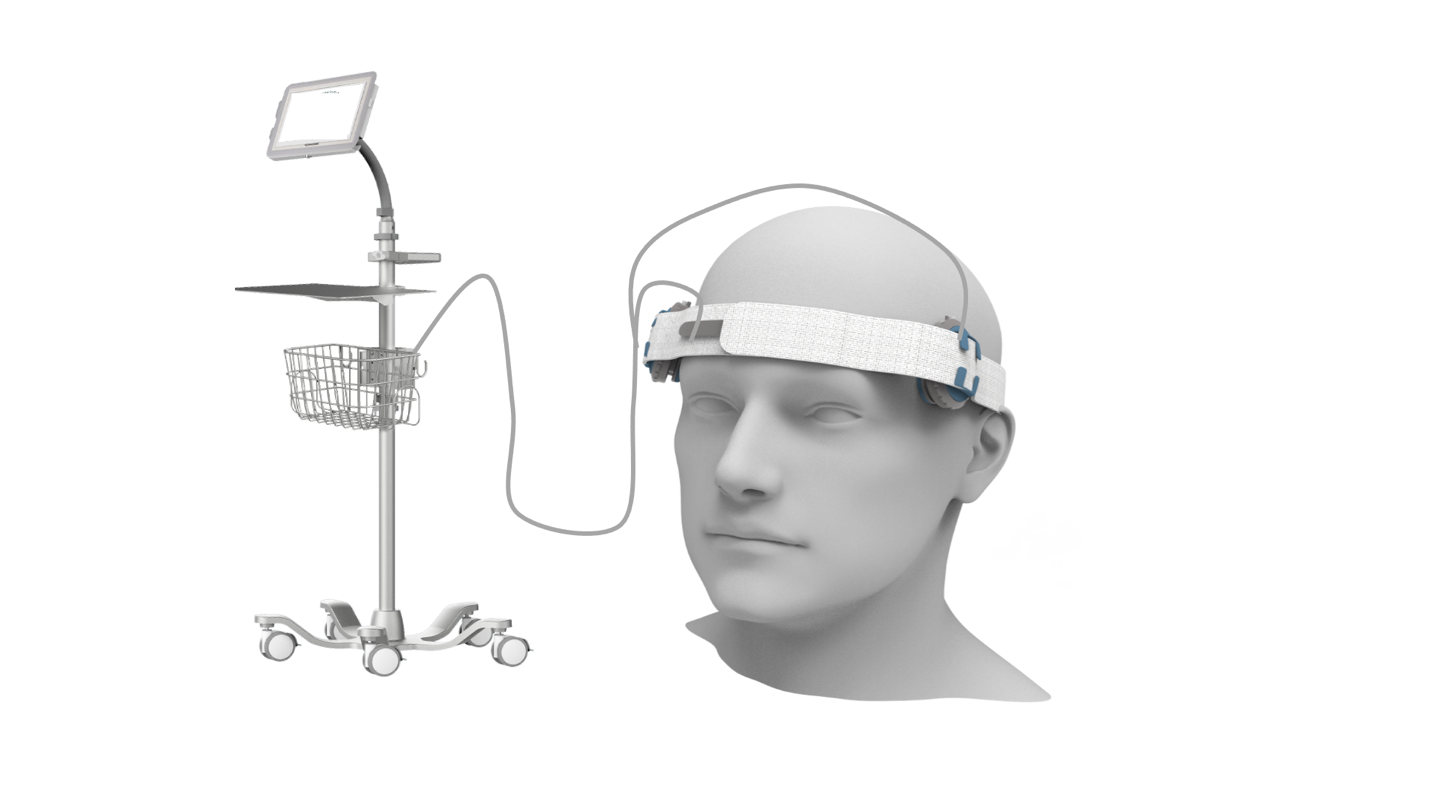

**Figure 3.** The Optical Brain Pulse Monitor

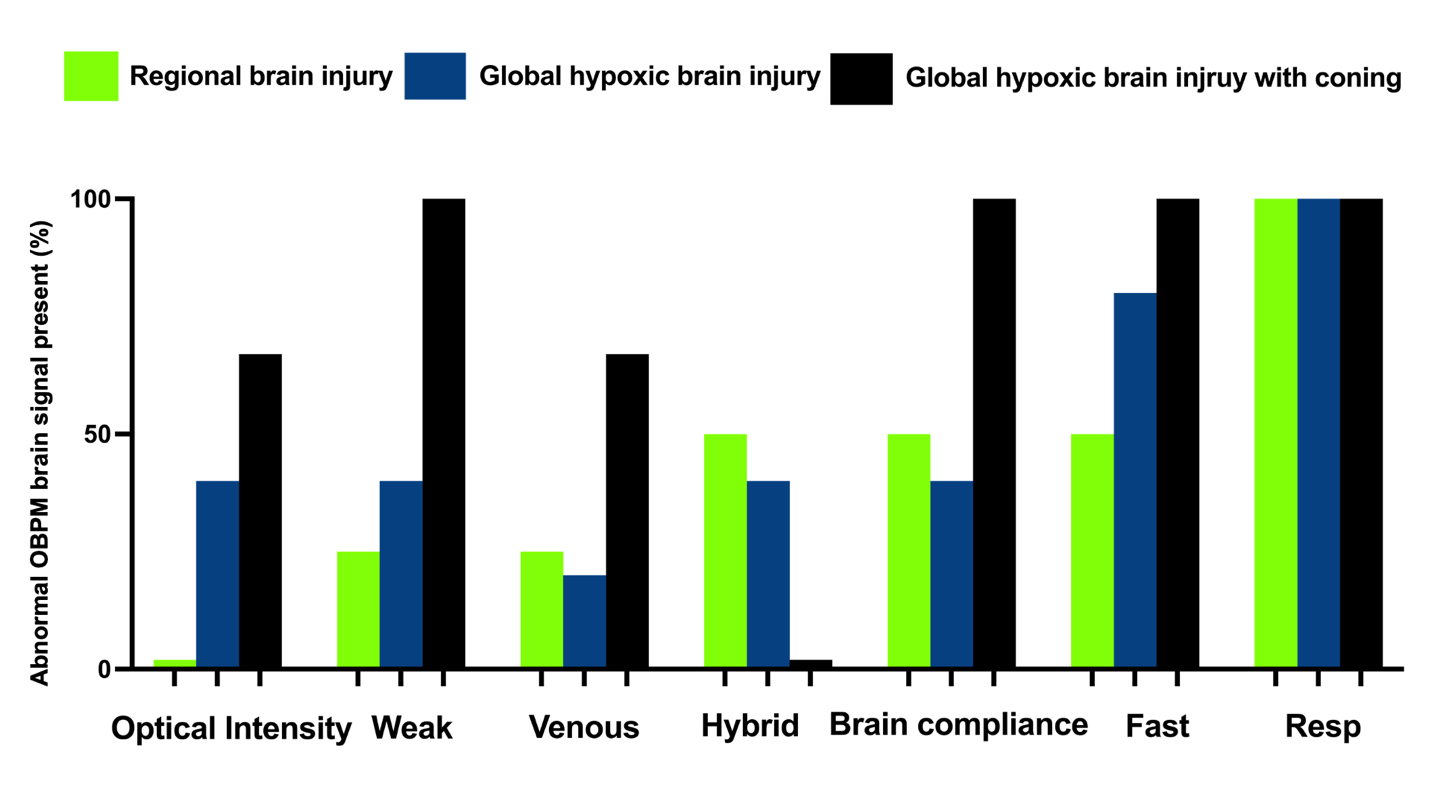

**Figure 4. Presence of abnormal types of OBPM signals in relation to the severity of brain injury.** There was variability in the OBPM abnormal signals. The lowest severity of brain injury was regional brain injury (green), global hypoxic-ischemic brain injury (blue) was more severe and global hypoxic brain injury with coning (black) was the severest. Abbreviations: **Weak**, Weak brain pulse; **Venous;** Venous I or II brain pulse, **Hybrid;** Hybrid brain pulse, **Brain compliance;** Low brain compliance pulse, **Fast;** Fast waves, **Resp**; High Respiratory wave amplitude.

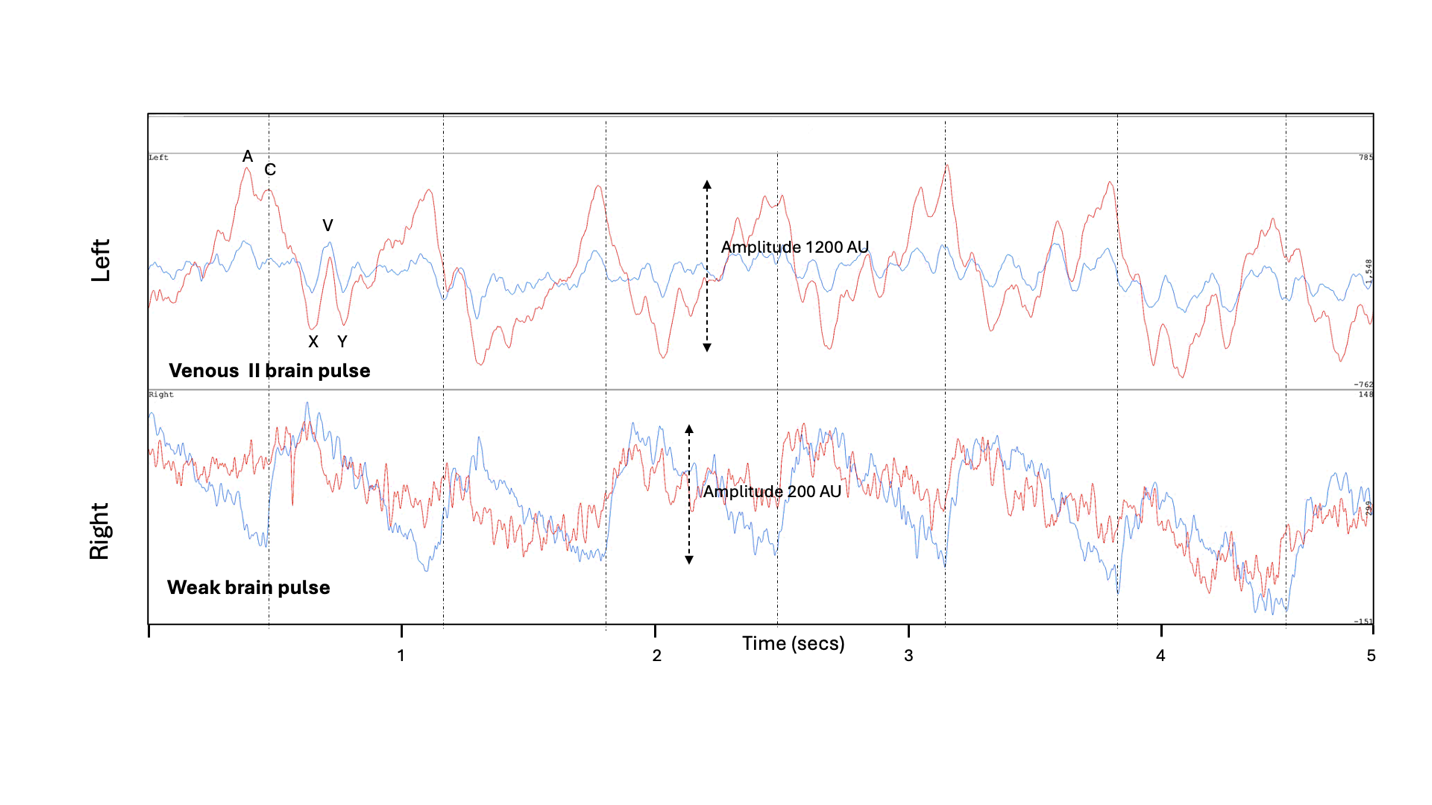
**Figure 5.** The left brain demonstrates a **Venous II brain pulse.** These features suggest low cerebral blood flow. The right brain demonstrates a **Weak brain pulse** which has a very low pulse amplitude, which is also associated with low cerebral blood flow. Red brain pulse is 940 nm and blue brain pulse 660 nm. *Abbreviations:* AU: arbitrary units

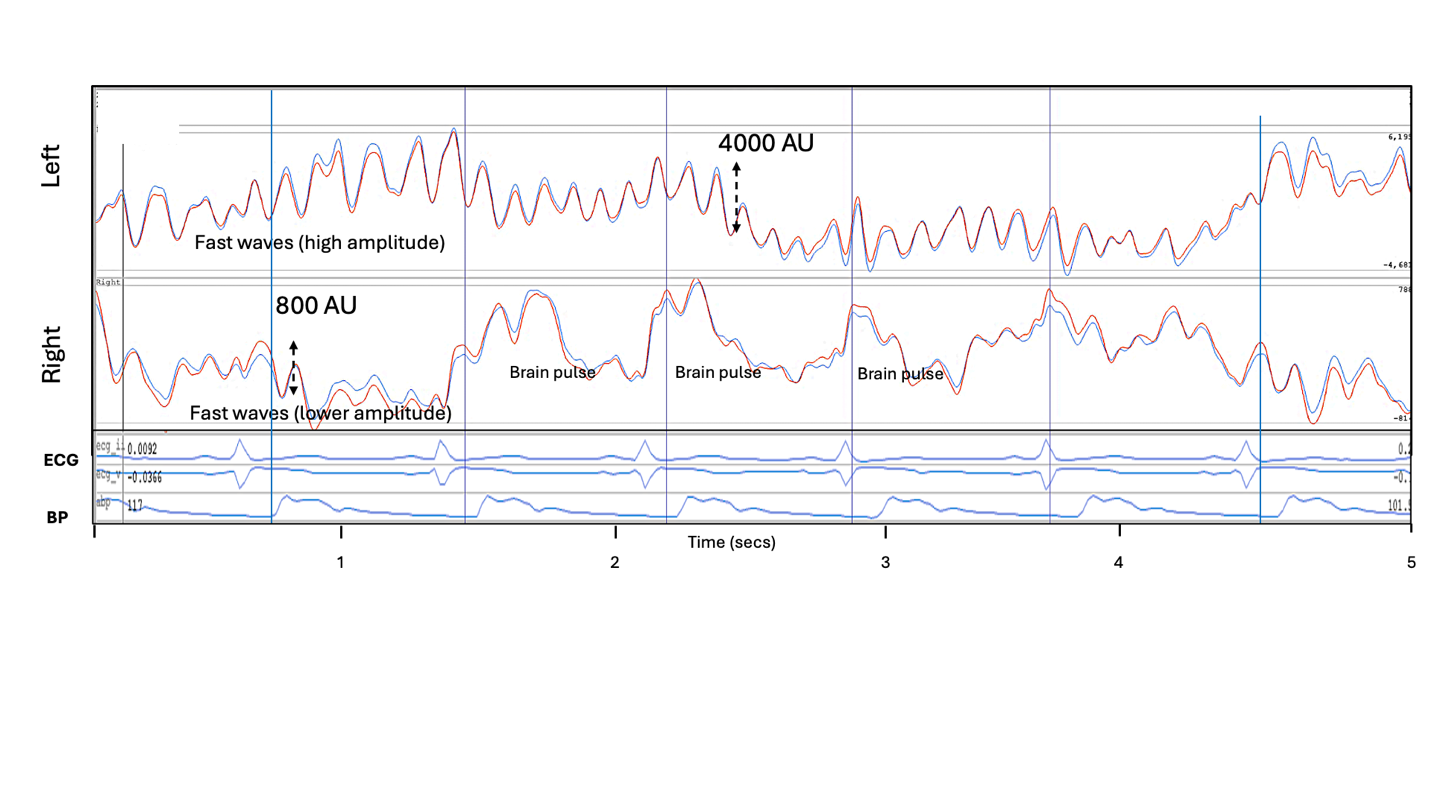

**Figure 6**. Left brain demonstrates **Fast waves** with high frequency oscillations (~ 8 Hz) of high amplitude (4000 AU). Over the right brain low amplitude (800 AU) **Fast waves are** combined with some cardiac pulses (blue vertical lines). Fast waves are a marker of the presence of a brain injury. If bilateral fast waves are present the side with the highest amplitude is likely the origin side. Fast waves may result from abnormally high amplitude fast oscillations of cerebrovascular smooth muscle with transmitted pressure waves through the brain tissue. Red brain pulse is 940 nm and blue brain pulse 660 nm. *Abbreviations* BP: blood pressure, ECG: electrocardiogram, AU: arbitrary units

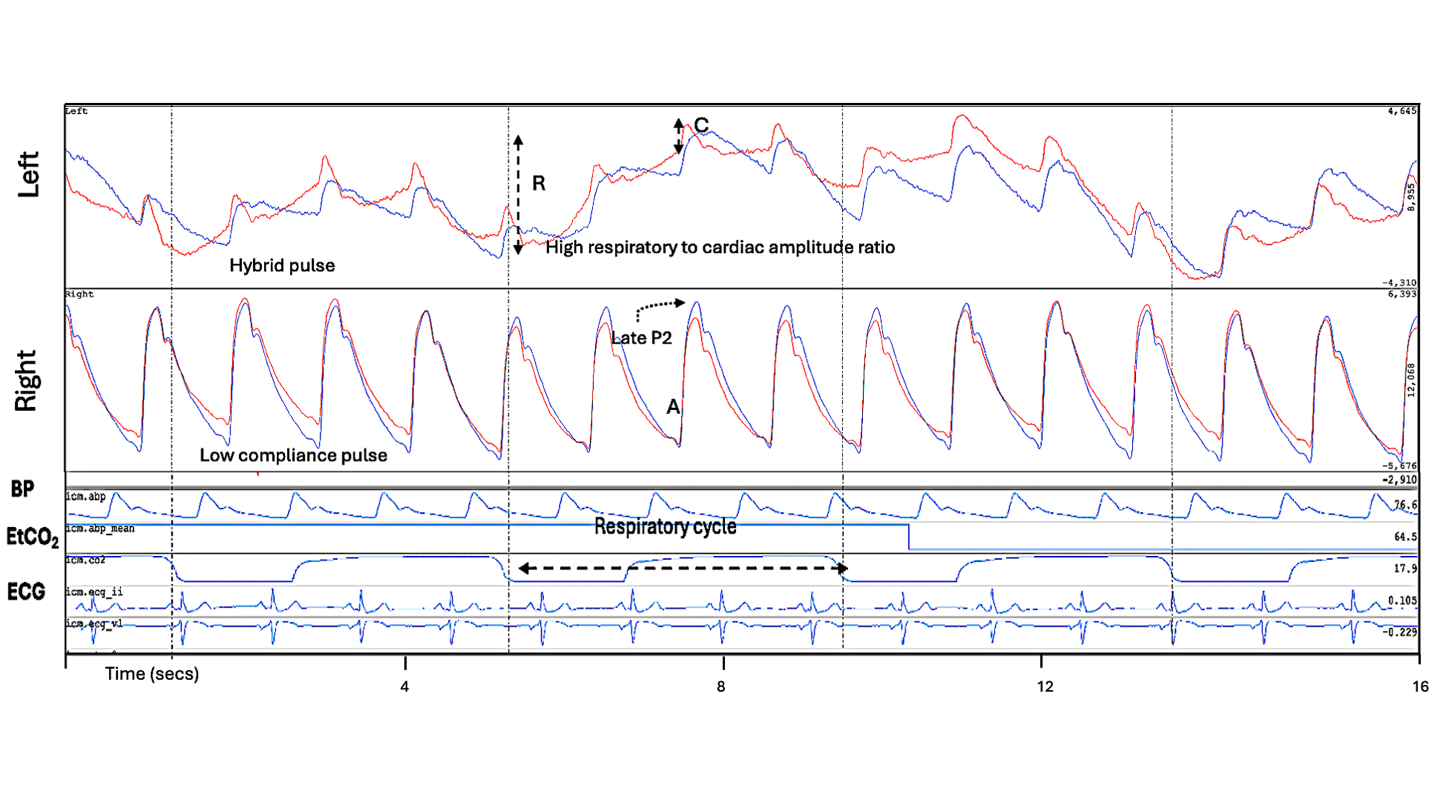

**Figure 7.** A patient following out of hospital cardiac arrest with a right posterior cerebral artery infarct on CT. Over the left brain **high respiratory amplitude waves** associated with **Hybrid brain pulses** are present. The right brain has a **Low compliance brain pulse,** demonstrating a late pulse peak (P2) and A wave is also present. These features suggest swelling or oedema of the right hemisphere with potential for mid-line shift of the brain to the left and low cerebral blood flow on the left hemisphere. Red brain pulse is 940 nm and blue brain pulse 660 nm. *Abbreviations* BP: blood pressure, EtCO_2_ : End-tidal CO_2_, ECG: electrocardiogram, R : respiratory wave amplitude, C : cardiac pulse amplitude; CT computerized tomography.

**Supplement Table 1: Characteristics associated with the Severity of brain injury**

|  | **Presentation** | **Invasive Treatments** | **Brain injury** | **Early detection assessable** | **WLST/Palliation** | **Monitoring days post injury** |
| --- | --- | --- | --- | --- | --- | --- |
| **Regional brain injury** | | |  |  |  |  |
| 1 | VT In hospital cardiac arrest 50 min ROSC | VA ECMO, CAGS | L MCA infarct (possibly late onset) on CT | Yes | Yes | 1,2 |
| 2 | VT OOH cardiac arrest 7 min ROSC | Coronary angiogram | R occipital infarct on CT.  Initial CT NAD | Yes | No, hospital transfer | 1,2 |
| 3 | VT OOH cardiac arrest 15 min ROSC | nil | L occipital infarct on CT.  Initial CT NAD | Yes | Yes | 2 |
| 4 | VT OOH cardiac arrest 28 min ROSC | LAD stent | R sided weakness, prolonged | Yes | No, hospital transfer | 1, 2 |
| **Global hypoxic ischemic brain injury** | | |  |  |  |  |
| 1 | PEA In hospital cardiac arrest 30 min ROSC | nil | GHII on EEG Initial CT NAD | Yes | Yes | 1,2,6 |
| 2 | Asystole OOH cardiac arrest 27 min ROSC | nil | GHII on CT | No | Yes | 1,2 |
| 3 | Asystole OOH cardiac arrest | OD pathway | GHII on CT | No | Yes | 3 |
| 4 | In hospital Respiratory arrest PEA 15 min ROSC | Coronary angiogram,  OD pathway | GHII on MRI  Initial CT NAD | Yes | Yes | 2,3,4 |
| 5 | SAH PEA OOH cardiac arrest ROSC 50 min | L craniectomy,  L PCOM coil,  Invasive ICP | GHII and L hemisphere infarcts on CT | No | Yes | 2, 3, 7 |
| **Global hypoxic ischemic brain injury**  **with coning** | | | | |  |  |
| 1 | SAH, PEA OOH cardiac arrest ROSC 29 mins | Invasive ICP  OD pathway | GHII on CT  Brain SPECT absent perfusion | No | Yes | 2,3,4 |
| 2 | PE OOH cardiac arrest PEA, ROSC 58 min | ECMO | GHII on CT | Yes | Yes | 0 |
| 3 | VT OOH cardiac arrest ROSC 25 min | ECMO,  LAD Stent | GHII on CT | Yes | Yes | 0,1,2 |

**Legend:** VT; ventricular tachycardia, L MCA; left middle cerebral artery, OOH; out of hospital, ROSC, return of spontaneous circulation, PEA; pulseless electrical activity, GHII: Global hypoxic ischemic injury, OD; organ donation, CT: computerized tomography, MRI: magnetic resonance imaging, L PCOM; left posterior communicating artery, LAD; left anterior descending artery, T-PA; tissue plasminogen activator, VA ECMO; venous arterial extra-corporeal membrane oxygenation, CAGS; coronary artery bypass grafting.
